# Sensitivity of dual-task motor performance to varying levels of cognitive impairment: a systematic review and quality assessment

**DOI:** 10.1101/2023.09.19.23295819

**Authors:** Maciej Kos, Misha Pavel, Holly B. Jimison, Jane S. Saczynski

## Abstract

Dementia is one of the key public challenges of this century, with the number of persons with dementia worldwide projected to reach 115 million by 2050. This review aimed to answer whether monitoring of motor performance alone and during a cognitively taxing task (dual-task) is sufficiently sensitive to distinguish between levels of cognitive function (normal function, mild cognitive impairment, dementia) and, thus, appropriate for dementia screening. In the reviewed 15 studies, cognitively healthy controls had a higher dual-task gait speed than persons with impaired cognition (9/12 studies). The difference between dual- and single-task gait speeds (dual-task cost) was lower in healthy controls (7/8 studies). Such differences were not detected between patients with mild cognitive impairment and Alzheimer’s disease.

These results suggest that monitoring of dual-task performance may be used in early dementia screening. Diversity in research designs, lack of established statistical and reporting standards prevent meta-analysis of data.

## Introduction

Nearly 46 million people worldwide suffered from some form of dementia in 2015, with this number estimated to increase by six to ten million new cases per year^1, 2^. In the US, alone, Alzheimer’s disease affected an estimated 5·4 million in 2017 and was the fifth leading cause of death among older Americans^3^. As the number of persons with dementia is projected to more than double by 2050, reaching 115 million worldwide^2^, dementia constitutes one of the major public health challenges of this century.

Since most diseases are more manageable if detected early and their symptoms are objectively measurable, it would be beneficial to find more sensitive methods for early detection and more reliable assessment.

Dementia is characterized by cognitive deficits but also manifests itself in a wide spectrum of physical symptoms, including motor dysfunctions and an increased risk of falling^4, 5^. Of critical importance and relevance to cognitive assessments is the growing body of research that points to dementia-resultant attention deterioration^6^ as a risk factor for falls, linking cognitive impairment with motor function^7^.

Evidence originating from dual-task studies shows empirically that the concurrent execution of multiple tasks, e.g., walking and talking at the same time, results in degraded performance on either or both tasks. In such studies, participants complete at least two types of tests – often, a single-task test and dual-task test – each with a different level of cognitive load (CL). For example, in the first test, subjects only walk (single-task test, low CL) while in the second test the same participants walk and say the alphabet backward at the same time (dual-task test, high CL). The difference in the outcome variable – here, walking speed – measured during the single- and dual-task tests is referred to as a dual-task cost (DTC). A frequent explanation for this phenomenon is that higher levels of CL limit attentional resources more than lower levels. DTC is assumed to allow researchers to quantify the impact of attention impairment on the measured outcome. Therefore, dual-task experiments may provide valuable insight into how declining attentional capacity is mirrored in motor function, enabling accurate modeling of this phenomenon.

While research on motor function and dementia is rich^8–14^, relatively few studies investigate this relationship in a dual-task paradigm. Heterogeneity in study designs, reported measures and implementations, as well as relatively simple statistical analysis and insufficient focus on test-retest reliability in DT studies, further inhibit our ability to synthesize findings and incorporate results into diagnosis and clinical care. Therefore, a systematic review of existing high-quality evidence aimed at determining whether dual-task cost is sensitive to detecting cognitive changes, by grouping findings by experimental design and outcome, is essential to synthesizing results, establishing best practices and guiding future research in this area.

Given the prevalence of dementia, it is critical to deepen our understanding of this condition using the most accurate and reliable of available methods. A precise characterization of its relationship with motor abnormalities will enable identification of initial symptoms and earlier detection of dementia, potentially providing sufficient time to intervene and delay its progress.

With this goal in mind, this review complements previous reviews^15, 16^ by systematically assessing the quality of existing research and answering whether the impact of attention impairment on motor function – operationalized as dual-task cost – differentiates patients with varying levels of cognitive function (normal function, MCI, dementia).

### Search strategy and selection criteria

To identify relevant evidence, the authors searched PubMed and Google Scholar databases in April of 2019 using the following queries: “motor AND dual task AND dementia, “Motor behavior AND dual task AND dementia.”

To be included in this review, studies needed to: 1) be peer-reviewed and published in English between January 1970 and April 2019, 2) use validated measures (e.g., the Mini-Mental State Examination^17^ or the Montreal Cognitive Assessment^18^), a neuropsychological exam or medical history to identify participants with cognitive impairment, 3) compare at least two groups with different levels of cognitive function, e.g., healthy controls vs. mild cognitive impairment or mild cognitive impairment vs. Alzheimer’s disease, and 4) provide a quantitative objective measurement of motor function during a dual-task test. Furthermore, the authors excluded studies on participants with conditions that could influence motor performance, e.g., traumatic brain injury or dyspraxia. Relevant studies cited in articles meeting eligibility criteria were added to the set of reviewed papers. Literature reviews, expert opinions, and commentaries were excluded from this review.

### Data Collection

Abstracts of all retrieved publications were assessed for relevance. The resultant subset of studies was reviewed to determine if inclusion criteria were met. The authors abstracted data published in those papers that met this requirement using a standardized form. The abstracted information included: study type (cross-sectional or longitudinal), how cognitive function was assessed, sample characteristics (size, inclusion and exclusion criteria, mean age, comorbidities), characteristics of comparison groups (e.g., how cognitive impairment was defined, type and severity of dementia, size, mean age), details of tests used (single-task, dual-task and how they were implemented), reported measures of motor function, and statistical findings relevant to the objective of this review.

### Quality Assessment

To assess the quality of each study, the authors used Downs and Black criteria^19^. Following previous research^20, 21^, the scale was adapted to match the nature of reviewed studies better. In particular, we removed nine items regarding follow-ups, adverse effects, blinding, compliance, and randomization of group assignment.

Furthermore, based on established practice^22^, the authors modified the question about statistical power. The revised quality assessment evaluates whether reviewed studies presented a power calculation including determination of the sample size necessary to detect relevant effect sizes (Yes/No). This revised instrument provides a score from 0 to 19 points (Appendix A) with higher values corresponding to studies of higher quality.

### Definitions of change and difference in motor function

We defined a change in motor function parameters as a statistically significant change (p ≤ 0·05) in at least one such parameter between the single- and dual-task performance measurements of the same group of subjects (within-subject design). For example, if the average difference in walking speeds between single- and dual-task (for instance, 3 m/s and 1 m/s, respectively) was 2 m/s (p=0·001), then this result is reported as a change in motor function. The dual-task cost in this example is 2 m/s (|3 m/s – 1 m/s|).

We define a difference in motor function between two groups of subjects to be statistically different if at least one parameter of motor function shows a significant change (p ≤ 0·05) between two groups of subjects during either single-task or dual-task (between-group design). For example, if mean walking speed during dual-task was 4 m/s for patients with dementia and 2 m/s for healthy controls (p=0·001), then this result is reported as a difference in motor function. If the duals-task cost is 3 m/s for patients with dementia and 1 m/s for healthy controls (p=0·001), then this result is reported as a dual-task cost difference.

## Results

### Study Selection

The initial search returned 224 titles. After removing 13 duplicates, 211 abstracts were reviewed for eligibility, of which 190 were subsequently excluded. The resulting set of 21 articles was reviewed fully. Additionally, the authors identified six more relevant articles in the bibliographies of studies under review. These articles also underwent full examination. In total, 27 articles were fully reviewed; the authors excluded 12 of them. Consequently, this systematic review is based on the remaining 15 studies (Figure 1).

**Figure 1.**
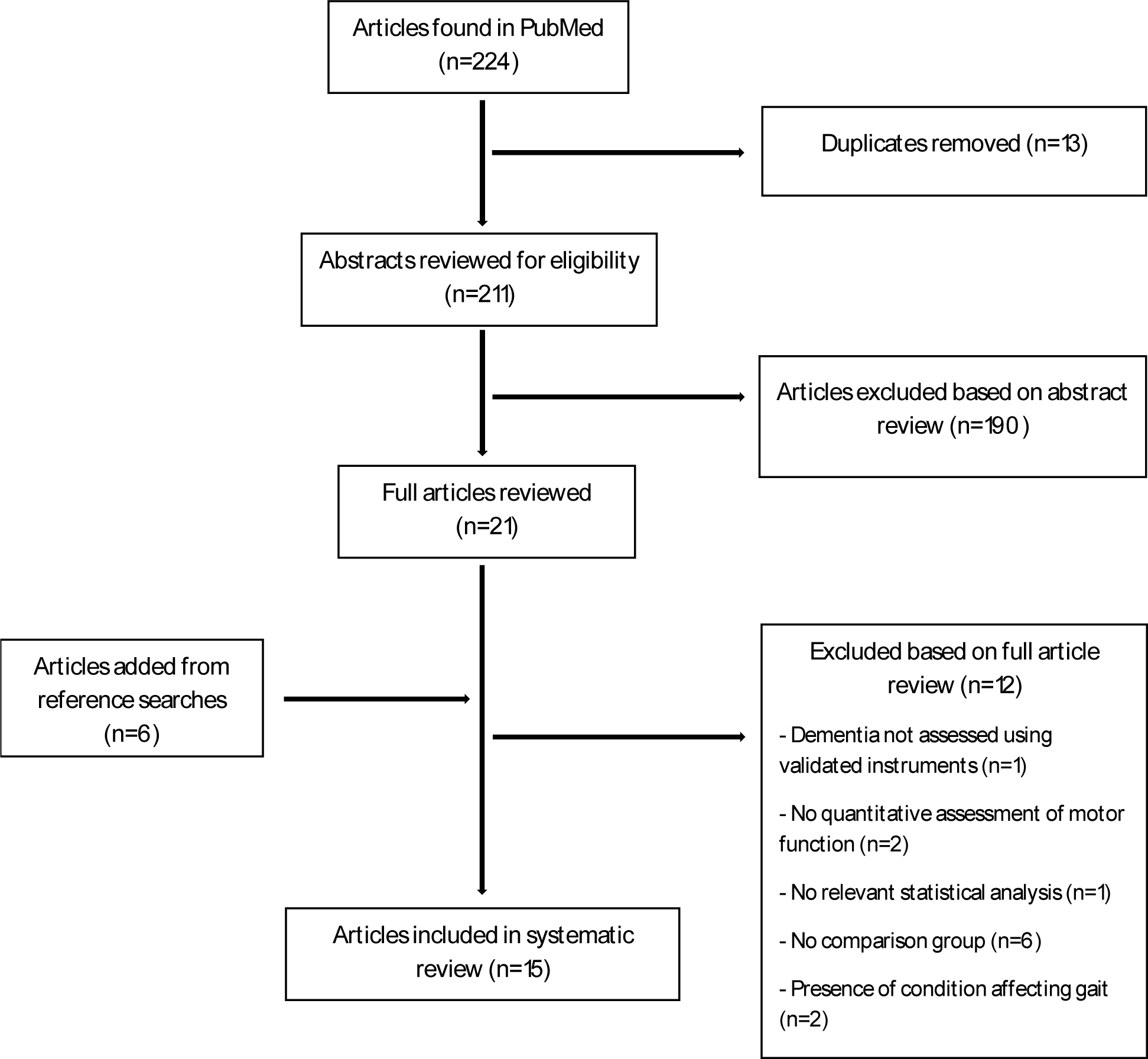
PRISMA-style flow diagram of the selection process.

### Study characteristics

All studies included in this review were cross-sectional. Ten^23–32^ of the studies focused solely on walking tasks. Three studies^33–35^ focused on only the “Timed Up and Go” task (TUG), which consists of not only walking but also of rising from an armchair. Two studies^36, 37^ included both TUG and walking tasks (Table 1).

**Table 1.**
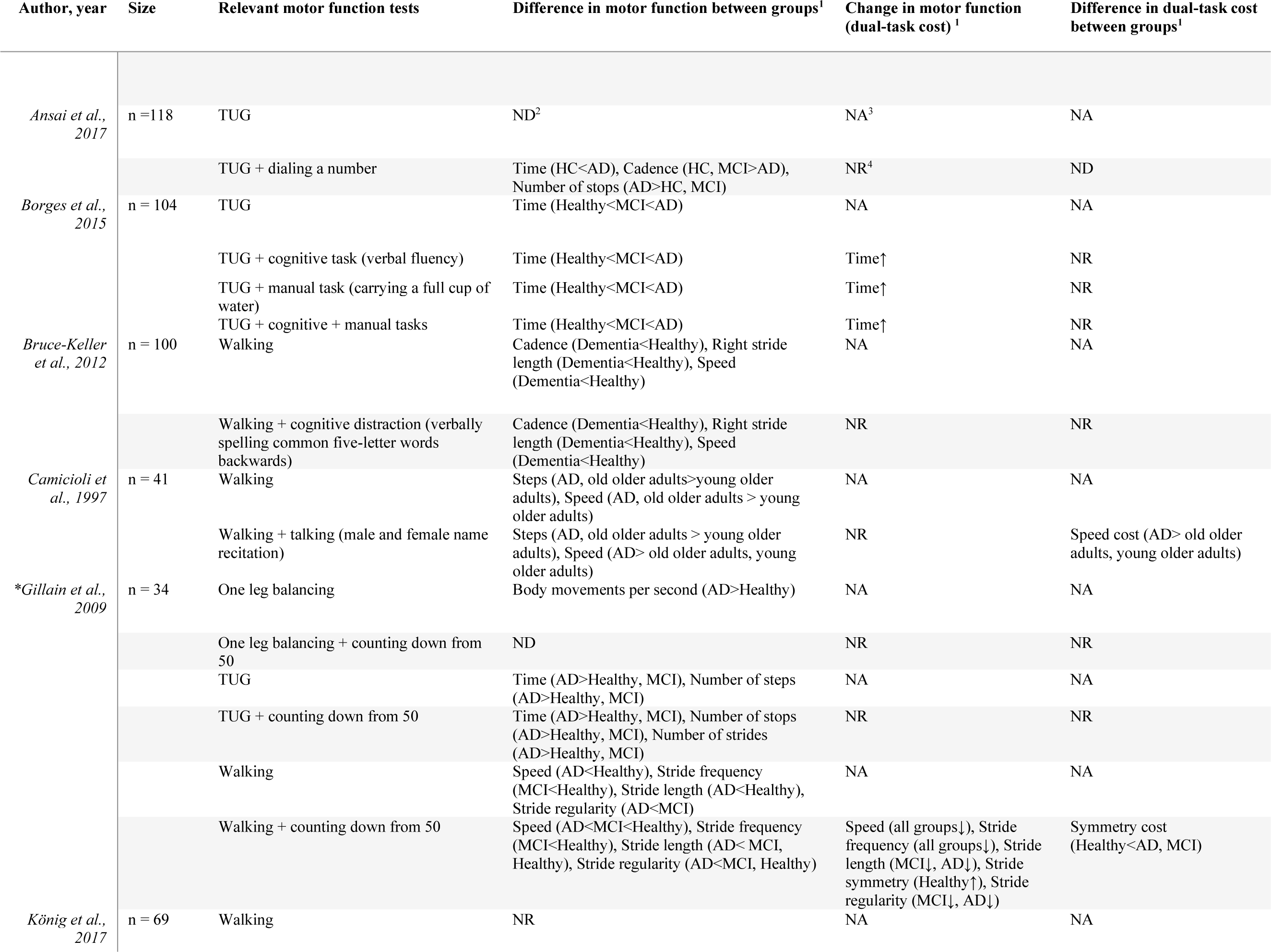
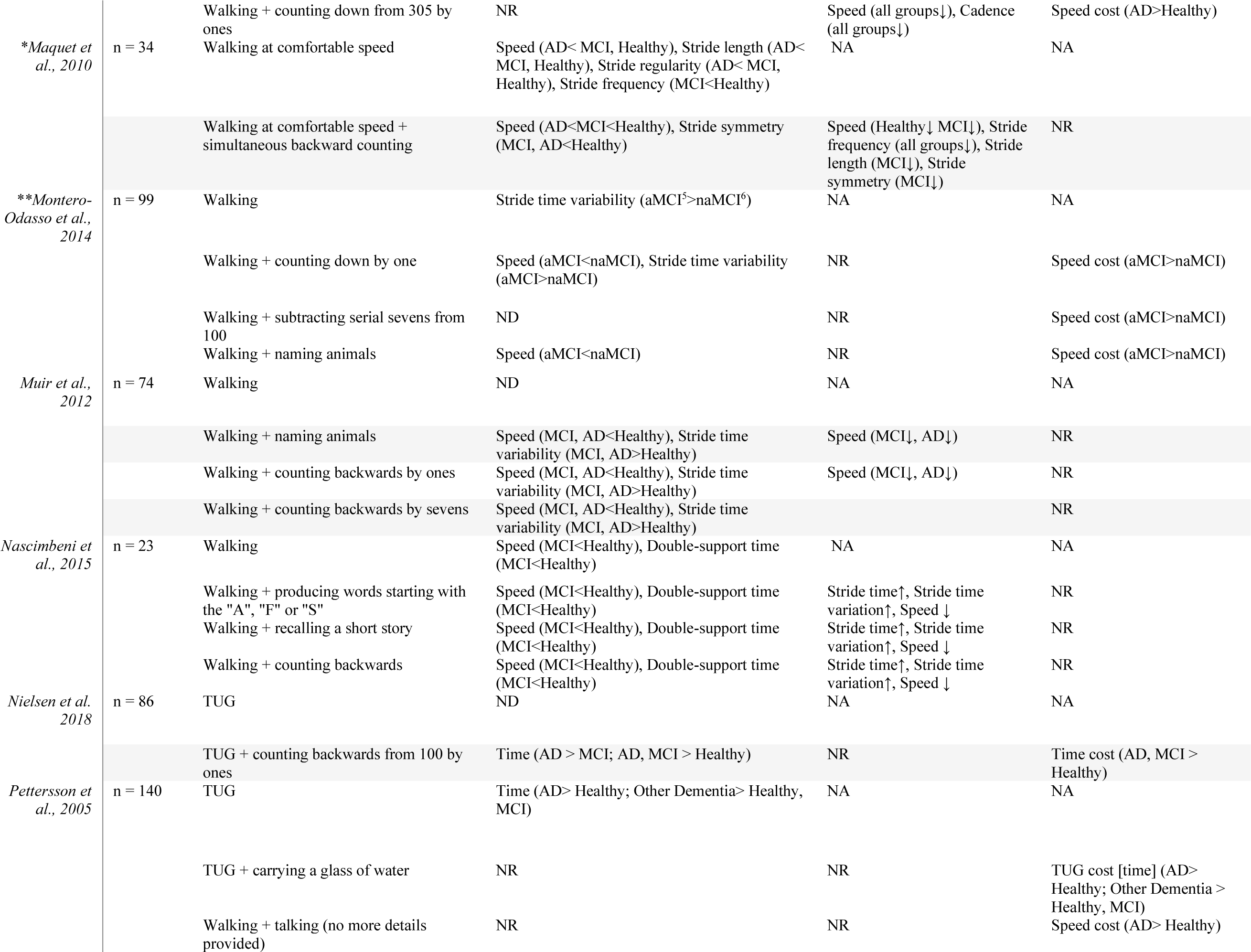
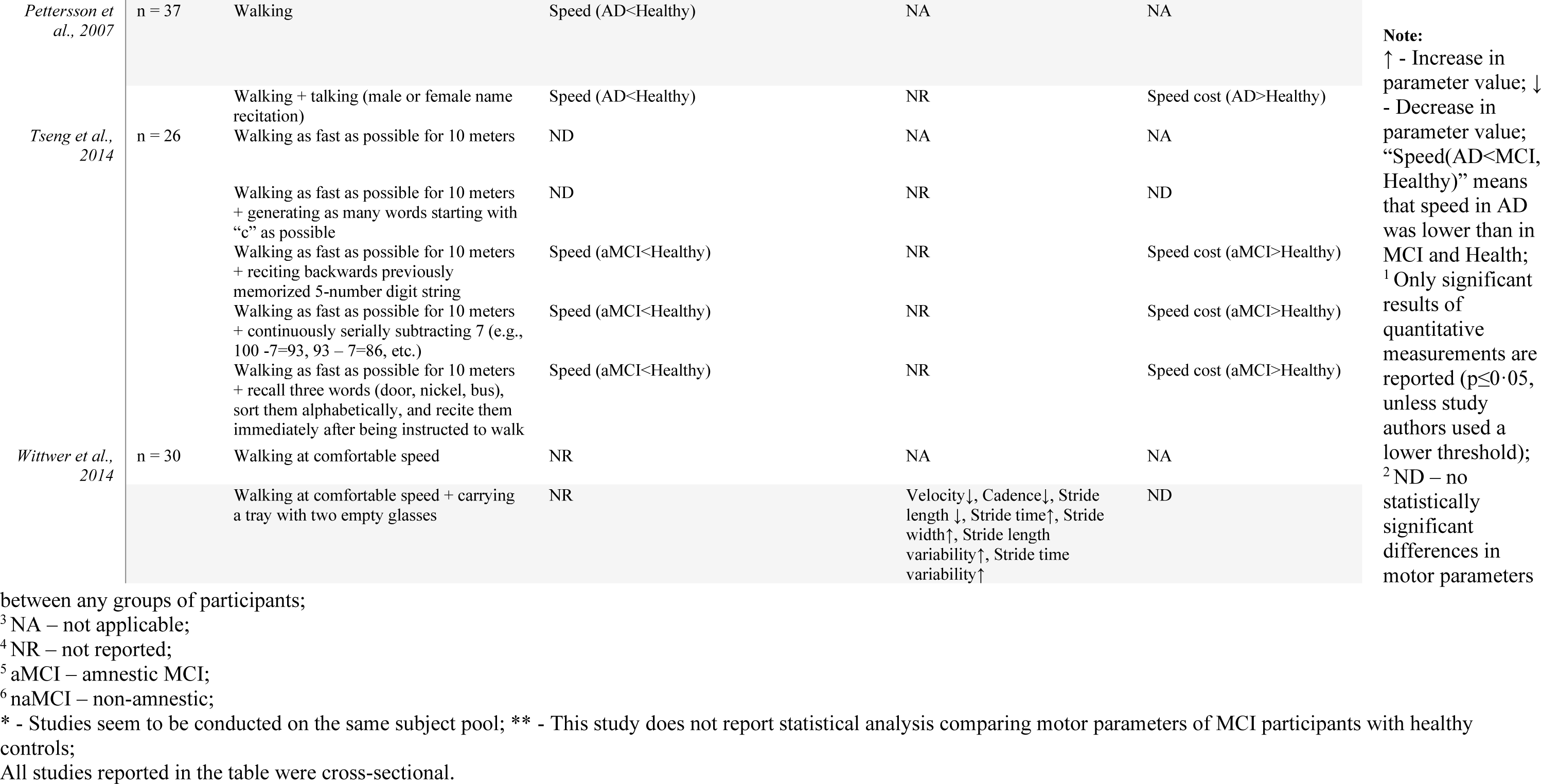
Table of evidence.

All included studies were lab experiments with two to four comparison groups (µ=3, σ=0·56). Mean group size per study was 24·31 (σ=13·77).

With one exception^31^, all studies included a control group of healthy participants. However, one study^38^ conducted on a sample of healthy controls did not include this group in its statistical characterization of motor parameters. Twelve studies^25–30, 33, 36, 37^ investigated patients with mild cognitive impairment (MCI); two of these articles^26, 30^ focused on amnestic MCI (aMCI) (Table 3). Ten studies^24, 25, 27, 29, 32–37^ included patients with Alzheimer’s, ^32, 34 35^disease (AD).

**Table 3.** Experimental groups.

| Author | Condition |  |  |  |  |  |  |  |  |  |
| --- | --- | --- | --- | --- | --- | --- | --- | --- | --- | --- |
|  | Healthy controls | AD | MCI | Amnesic MCI | Non-amnesic MCI | Mild AD | Dementia | Moderate to severe dementia | Mild dementia | Other dementia |
| *Ansai et al., 2017* | ✓ |  | ✓ |  |  | ✓ |  |  |  |  |
| *Borges et al., 2015* | ✓ | ✓ | ✓ |  |  |  |  |  |  |  |
| *Bruce-Keller et al., 2012* | ✓ |  |  |  |  |  | ✓ |  |  |  |
| *Camicioli et al., 1997* | ✓ | ✓ |  |  |  |  |  |  |  |  |
| *\*Gillain et al., 2009* | ✓ | ✓ | ✓ |  |  |  |  |  |  |  |
| *König et al. 2017* | ✓ | ✓ | ✓ |  |  |  |  |  |  |  |
| *\*Maquet et al., 2010* | ✓ | ✓ | ✓ |  |  |  |  |  |  |  |
| *\*\*Montero-Odasso et al., 2014* | ✓ |  | ✓ | ✓ | ✓ |  |  |  |  |  |
| *Muir et al., 2012* | ✓ | ✓ | ✓ |  |  |  |  |  |  |  |
| *Nascimbeni et al., 2015* | ✓ |  | ✓ |  |  |  |  |  |  |  |
| *Nielsen et al. 2018* | ✓ | ✓ | ✓ |  |  |  |  |  |  |  |
| *Pettersson et al., 2007* | ✓ | ✓ | ✓ |  |  |  |  |  |  |  |
| *Pettersson, 2005* | ✓ | ✓ | ✓ |  |  |  |  |  |  | ✓ |
| *Tseng et al., 2014* | ✓ |  | ✓ | ✓ |  |  |  |  |  |  |
| *Wittwer et al., 2014* |  |  |  |  |  |  |  | ✓ | ✓ |  |
| *sum* | 14 | 9 | 12 | 2 | 1 | 1 | 1 | 1 | 1 | 1 |
| *%* | 93.33% | 60.00% | 80.00% | 13.33% | 6.67% | 6.67% | 6.67% | 6.67% | 6.67% | 6.67% |

\* Studies seem to be conducted on the same subject pool; \*\* This study does not report statistical analysis comparing motor parameters of MCI participants with healthy controls

### Assessment of cognitive function

All studies except one^28^ used Mini-Mental State Examination^17^ (MMSE) as a quantitative criterion for group assignment. Six studies^23–25, 30, 34, 37^ used Clinical Dementia Rating^39^ (CDR), two used^26, 27^ Montreal Cognitive Assessment^18^ (MoCA), and one^28^ used Milan Overall Dementia Assessment^40^ (MODA).

### Participant characteristics

Most participants were older adults. Mean sample ages ranged from 55±4·7^29^ to 80·5±5·9^31^. None of the studies provided information about participants’ socio-economic status (however, six studies^23, 27, 30, 33–35^ did report participants’ education). Only one study included information about ethnicity^24^.

### Quality assessment

The quality of included studies was assessed using modified Downs and Black criteria^19^ (Appendix A). Scores ranged from eight to 14 (42% to 73·5%) out of 19 with a mean of 11 and a standard deviation of 1·69 (57·9% and 8·9%, respectively). Only one study^34^ presented calculations of power to determine the appropriate sample size. None of the studies demonstrated that participants were representative of entire relevant populations with a similar health status (Downs and Black criteria^19^, Appendix A: question 10.).

### Reported gait parameters in dual-task studies

Overall, 15 different gait parameters were reported in 12 articles that included a DT walking task (Table 3). All studies reported gait speed^23–32, 36, 37^, four reported cadence^23, 24, 31, 32^ and stride time variability^26–28, 31^. Six parameters were reported only once. On average, studies that scored over 50% on quality assessment reported fewer gait parameters than those that scored 50% or lower (2·7 vs. 6, respectively).

### Motor function

#### Between-group differences in dual-task performance

##### GAIT

During dual-task tests, cognitively impaired individuals walked slower than healthy controls and their stride time variability was higher. Nine^23–30, 37^ of the 12 studies^23–32, 36, 37^ reported between-groups differences in gait speed during dual-task tests. Two^26, 27^ out of four studies^26–28, 31^ reported similar differences in stride time variability (Tables 4, 5).

**Table 4.** Gait parameters in dual-task tests.

| Author, year | Speed | Cadence | Stride time variability | Stops | Stride length | Stride frequency | Stride regularity | Stride symmetry | Stride time | Double support time | Right stride length | Step number | Step time variance | Stride length variability | Stride width |
| --- | --- | --- | --- | --- | --- | --- | --- | --- | --- | --- | --- | --- | --- | --- | --- |
| *Bruce-Keller et al., 2012* | ✓<sup>D</sup> | ✓<sup>D</sup> |  |  |  |  |  |  |  |  | ✓<sup>D</sup> |  |  |  |  |
| *Camicioli et al., 1997\** | ✓<sup>D, DTCd</sup> | ✓ |  |  |  |  |  |  |  |  |  | ✓<sup>D</sup> |  |  |  |
| *\*Gillain et al., 2009* | ✓<sup>D, DTC</sup> |  |  | ✓ | ✓<sup>D, DTC</sup> | ✓<sup>D, DTC</sup> | ✓<sup>D, DTC</sup> | ✓<sup>DTC, DTCd</sup> |  |  |  |  |  |  |  |
| *König et al., 2017* | ✓<sup>DTC, DTCd</sup> | ✓<sup>DTC</sup> |  |  |  |  |  |  |  |  |  |  | ✓ |  |  |
| *\*Maquet et al., 2010* | ✓<sup>D, DTC</sup> |  |  | ✓ | ✓<sup>DTC</sup> | ✓<sup>D, DTC</sup> | ✓ | ✓<sup>D, DTC</sup> |  |  |  |  |  |  |  |
| *Montero-Odasso et al., 2014* | ✓<sup>D, DTCd</sup> |  | ✓<sup>D</sup> |  |  |  |  |  |  |  |  |  |  |  |  |
| *Muir et al., 2012* | ✓<sup>D, DTC</sup> |  | ✓<sup>D</sup> |  |  |  |  |  |  |  |  |  |  |  |  |
| *Nascimbeni et al., 2015* | ✓<sup>D, DTC</sup> |  | ✓<sup>DTC</sup> |  |  |  |  |  | ✓<sup>DTC</sup> | ✓<sup>D</sup> |  |  |  |  |  |
| *Pettersson, 2005* | ✓<sup>DTCd</sup> |  |  |  |  |  |  |  |  |  |  |  |  |  |  |
| *Pettersson et al., 2007* | ✓<sup>D, DTCd</sup> |  |  |  |  |  |  |  |  |  |  |  |  |  |  |
| *Tseng et al., 2014* | ✓<sup>D, DTCd</sup> |  |  |  |  |  |  |  |  |  |  |  |  |  |  |
| *Wittwer et al., 2014* | ✓ | ✓ | ✓ |  | ✓ |  |  |  | ✓ |  |  |  |  | ✓ | ✓ |

**Note:**
\* - Studies conducted on the same subject pool;
✓ - This parameter was reported in this study (regardless of its statistical significance);
D – Dual-task difference: statistically significant difference in this parameter between at least two groups performing a dual-task ( $p \leq 0.05$ );
DTC – Dual-task cost: statistically significant change in this parameter between single and dual-task ( $p \leq 0.05$ );
DTCd – Dual-task cost difference: statistically significant difference in dual-task cost between at least two groups ( $p \leq 0.05$ );

**Table 5.** Number of studies reporting significant differences in DT performance and DTC.

| *Between-groups differences in:* | Walking tasks |  |  | TUG |  |  |
| --- | --- | --- | --- | --- | --- | --- |
|  | *Speed* | *Stride time variability* | *Stride symmetry* | *Time* | *Number of stops* | *Cadence* |
|  | *higher in healthy controls* | *lower in healthy controls* | *higher in healthy controls* | *shorter in healthy controls* | *lower in healthy controls* | *higher in healthy controls* |
| *dual-task performance* | 9 | 1 | 0 | 4 | 2 | 1 |
| *dual-task cost* | *Speed cost* | *Stride time variability cost* | *Stride symmetry cost* | *Time cost* | *Number of stops cost* | *Cadence cost* |
|  | *lower in healthy controls* |  |  |  |  |  |
|  | 7 | 0 | 1 | 2 | 0 | 0 |

##### TUG

Time to completion tends to be longer and the number of stops higher among participants with dementia than among healthy controls. Four^33–35, 37^ out of five TUG papers ^33–37^ reported between-group differences during dual-task tests. All four studies reported differences in time to completion; two in the number of stops^34, 37^, one in cadence^34^, and one in the number of strides^37^ (Tables 4, 5).

#### Between-groups differences in dual-task cost

##### GAIT

As a result of adding a secondary task, participants with cognitive impairments (AD, amnestic MCI) tend to slow down more than controls. Among 12 studies characterizing gait^23–32, 36, 37^, eight analyzed dual-task costs differences^24, 26, 29–32, 36, 37^; seven of them reported dual-task costs differences to be statistically significant^24, 26, 29, 30, 32, 36, 37^. In six^24, 26, 29, 30, 32, 36^ of the seven studies these results concerned gait speed; in the remaining one^37^, the difference regarded stride symmetry with healthy controls having a more symmetric stride than cognitively impaired participants (Tables 4, 5).

Only one^26^ of the seven studies reported a significant difference between groups of participants with varying levels of cognitive impairment; speed cost was higher for amnestic than for non-amnestic MCI participants. The remaining six studies^24, 29, 30, 32, 36, 37^ found healthy controls to walk faster than patients with cognitive impairment but were unable to differentiate between participants with AD and amnestic MCI (Tables 1, 4).

##### TUG

Among the five studies that included TUG task^33–37^, three analyzed differences in dual-task costs^34–36^. In two instances^35, 36^ time to completion increased statistically significantly more among participants with dementia than among controls (Table 5).

## Discussion

This systematic review provides robust evidence for differences in gait speed and dual-task cost for gait speed between healthy participants and individuals with cognitive impairment (MCI and AD) during dual-task tests. However, existing findings suggest such a difference is hard to detect between patients with MCI and AD. There is insufficient evidence to make any meaningful conclusions about other gait parameters and performance on the TUG test.

Identification of strong evidence for differences in dual-task cost for gait speed between healthy participants and individuals with cognitive impairment (MCI and AD), but not between participants with MCI and AD, as well as rigorous quality assessment, constitute novel contributions of this review. The second result, i.e., correlation between the severity of cognitive impairment and decreased motor performance under single- and dual-task conditions, confirms the conclusions of other reviews^15, 16, 41^ using systematically reviewed evidence from more contemporary dual-task studies focused on juxtaposing motor parameters from groups of participants with different levels of cognitive impairment.

The results suggest that simple dual-task motor function tests (e.g., walking and talking) might augment traditional screening tests for cognitive impairment^42^, especially if conducted regularly, for instance, during well-care visits. Furthermore, given the pervasiveness of falls in patients with dementia^43^, clinical practitioners should consider regularly screening patients at an elevated risk of fall complications (e.g., older adults with osteopenia, osteoporosis, and coagulation disorders) for changes in both cognitive decline and motor function.

We have identified several methodological issues in the literature. Reviewed studies reported 16 different gait parameters in total, of which only gait speed was reported in all articles. While the exploration of relationships between cognitive impairment and novel motor function parameters can lead to important discoveries, it is essential that a standardized set of motor metrics and statistical comparisons (e.g., between-groups differences in dual-task performance and in dual-task cost) is consistently reported in all articles, even in the absence of statistical significance. Such harmonization would enable easier comparison of results across studies as well as facilitate evaluation, generalization, translation, and meta-analysis of findings.

Reviewed studies employed a diverse set of cognitive tasks in their novel implementations of the dual-task framework. We recognize that these new designs are frequently well-justified. However, to facilitate results comparison, they should augment established methods, but not replace them. The Canadian Consortium on Neurodegeneration in Aging provides excellent guidelines for the standardization of gait assessments^44^.

To ensure that results are generalizable, it is crucial to demonstrate the representativeness of participants and recruitment facilities. Unfortunately, none of the reviewed studies showed that recruited subject pools share characteristics with the older adult population, which could have been achieved by demonstrating that the main confounders in the participant pool follow the same distributions as in the reference population (Downs and Black criteria^19^, Appendix A: question 10.). Given a high prevalence of depression among AD patients^45^, the exclusion of depressed patients from some of the reviewed studies suggests that generalizability of findings may be limited. Furthermore, none of the reviewed articles reported information about the participants’ socioeconomic status (SES). In the light of very strong evidence for the impact of SES on health outcomes^46, 47^ stemming from, e.g., economic (e.g., limited access to healthcare) or biological (e.g., brain’s different levels of exposure to stress over lifetime^48^) reasons, it is crucial that researchers report, if not control for, SES, and conduct their studies on diverse populations.

Finally, only one study^34^ presented evidence that sample sizes were determined using power calculations. Consequently, interpretation of reported findings, particularly involving samples of fewer than ten participants, is challenging, even if such studies were otherwise well-designed.

The Canadian Consortium’s on Neurodegeneration in Aging gait assessment guidelines^44^ provide a strong foundation for harmonization of dual-task research. Furthering this goal requires the development of a standardized set of statistical comparisons to be reported in all studies. For instance, it is possible that although within-group changes in walking speed from a single-task to a dual-task are not statistically significant, the difference between the groups in the dual-task cost is. The correct interpretation of the latter result when the former one is not reported might be challenging. One can envision future guidelines to include a rubric outlining a set of required statistical analyses thereby further increasing reporting quality and accelerating scientific progress.

The primary focus of reviewed studies was on improving our understanding of the interplay of cognition and motor function. Currently, the dominating approach in this strain of research is testing if - and under what conditions - the motor function of groups of healthy individuals and patients with varying levels of cognitive impairment differ from one another. However, existing strong evidence for the presence of such group differences does not guarantee clinical efficacy of motor function testing in detecting dementia. Established data-driven techniques and computational models^49^ could enhance traditional statistical used in emerging studies in this space by allowing to evaluate the diagnostic performance of such tests using longitudinal patient-level (idiographic), not cohort-level (nomothetic), data. Furthermore, examination of the trade-offs between cognitive and motor performance with varying relative utility of each task may further deepen our understanding of the underlying conditions. Such analysis could potentially improve our interpretation of the current metrics such the dual-task cost.

All studies in this review relied on gait assessments conducted in laboratory settings. New research using data collected *in situ* is needed to assess the ecological validity of existing findings and aid patients at risk of cognitive impairment. The advent of wearable monitoring devices and other unobtrusive remote sensors^50, 51^ allows for continuous collection of temporary dense human motor data opening a new avenue for research that may enable early detection of dementia^49^.

## Conclusions

Healthy and cognitively impaired individuals differ in gait speed and changes in gait speed when performing dual-task tests. However, existing evidence suggests no differences in dual-task costs between patients with MCI and AD. Harmonization of reported motor parameters and comparative statistical tests, as well as ensuring that recruited samples are representative and of sufficient size will strengthen the quality of evidence. Applying modern analytical approaches to patient-level, ecologically valid data will improve the clinical relevance of research in this area.

## Data Availability

No data are available. This is a systematic literature review.

## Acknowledgments

MK was partially funded by the Association for Computing Machinery/Intel Corporation Computational and Data Science Fellowship as well as by Google Scholarship. Neither of these organization had any involvement in this study.

## Contributions

Conceptualization of the study: MK, JSS, MP; Literature search, abstract screening, article review: MK; Figures, Tables: MK, JSS; Writing: MK, JSS; Revisions for intellectual content: MK, JSS, HBJ, MP

## Appendix A. Quality assessment form

|  |  |
| --- | --- |
| 1. Is the hypothesis/aim/objective of the study clearly described? | Yes = 1;<br>No = 0 |
| **2. Are the main outcomes to be measured clearly described in the introduction or methods section?** If the main outcomes are first mentioned in the Results section, the question should be answered no. | Yes = 1;<br>No = 0 |
| **3. Are the characteristics of the patients included in the study clearly described?** In cohort studies and trials, inclusion and/or exclusion criteria should be reported. In case-control studies, a case-definition and the source for controls should be provided. | Yes = 1;<br>No = 0 |
| **4. Are the interventions of interest clearly described?** Treatments and placebo (where relevant) that are to be compared should be clearly described. | Yes = 1;<br>No = 0 |
| **5. Are the distributions of principal confounders in each group of patients to be compared clearly described?** A list of principal confounders is provided. | Yes = 2;<br>Partially = 1;<br>No = 0 |
| **6. Are the main findings of the study clearly described?** Simple outcome data (including denominators and numerators) should be reported for all major findings so that the reader can check the major analyses and conclusions. (This question does not cover statistical tests that are considered below). | Yes = 1;<br>No = 0 |
| **7. Does the study provide estimates of the random variability in the data for the main outcomes?**<br>In non-normally distributed data, the inter-quartile range of results should be reported. In normally distributed data, the standard error, standard deviation, or confidence intervals should be reported. If the distribution of the data is not described, it must be assumed that the estimates used were appropriate and the question should be answered yes. | Yes = 1;<br>No = 0 |
| **8. Have actual probability values been reported (e.g., 0.035 rather than <0.05) for the main outcomes, except where the probability value is <0.001?**<br>If no *p* values are presented, the question should be answered 'no'. If *p* values presented and there is a mixture of reporting (some presented as < or > specific figures, some as equality, e.g. *p* = 0.034), question should be answered 'yes'. | Yes = 1;<br>No = 0 |
| **9. Were the subjects asked to participate in the study representative of the entire population from which they were recruited?**<br>The study must identify the source population for patients and describe how the patients were selected. Patients would be representative if they comprised the entire source population, an unselected sample of consecutive patients, or a random sample. Random sampling is only feasible where a list of all members of the relevant population exists.<br>Where a study does not report the proportion of the source population from which the patients are derived, the question should be answered as unable to determine. | Yes = 1;<br>No = 0;<br>Unable to determine = 0 |
| **10. Were those subjects who were prepared to participate representative of the entire population from which they were recruited?**<br>The proportion of those asked and agreed to participate should be stated.<br>Validation that the sample was representative would include demonstrating that the distribution of the main confounding factors was the same in the study sample and the source population. | Yes = 1;<br>No = 0;<br>Unable to determine = 0 |
| **11. Were the staff, places, and facilities where the patients were treated representative of the treatment the majority of patients receive?**<br>For the question to be answered 'yes' the study should demonstrate that the intervention was representative of that in use in the source population. The question should be answered no if, for example, the intervention was undertaken in a specialist center unrepresentative of the hospitals most of the source population would attend. | Yes = 1;<br>No = 0;<br>Unable to determine = 0 |
| **12. If any of the results of the study were based on "data dredging," was this made clear?**<br>Any analyses that had not been planned at the outset of the study should be clearly indicated. If no retrospective unplanned subgroup analyses were reported, then answer 'yes'. | Yes = 1;<br>No = 0;<br>Unable to determine = 0 |
| **13. Were the statistical tests used to assess the main outcomes appropriate?**<br>The statistical techniques used must be appropriate to the data. For example, nonparametric methods should be used for small sample sizes.<br>Where little statistical analysis has been undertaken but where there is no evidence of bias, the question should be answered yes. If the distribution of the data (normal or not) is not described it must be assumed that the estimates used were appropriate and the question should be answered 'yes'. | Yes = 1;<br>No = 0;<br>Unable to determine = 0 |
| **14. Were the main outcome measures used accurate (valid and reliable)?**<br>For studies where the outcome measures are clearly described, the question should be answered yes. For studies that refer to other work or demonstrates the outcome measures are accurate, the question should be answered as yes. | Yes = 1;<br>No = 0;<br>Unable to determine = 0 |
| **15. Were the patients in different intervention groups (trials and cohort studies) or were the cases and controls (case-control studies) recruited from the same population?**<br>For example, patients for all comparison groups should be selected from the same hospital. The question should be answered “unable to determine” for cohort and case control studies where there is no information concerning the source of patients included in the study. | Yes = 1;<br>No = 0;<br>Unable to determine = 0 |
| **16. Were study subjects in different intervention groups (trials and cohort studies) or were the cases and controls (case-control studies) recruited over the same period of time?**<br>For a study which does not specify the time period over which patients were recruited, the question should be answered as “unable to determine.” | Yes = 1;<br>No = 0;<br>Unable to determine = 0 |
| **17. Was there adequate adjustment for confounding in the analyses from which the main findings were drawn?**<br>This question should be answered no for trials if: the main conclusions of the study were based on analyses of treatment rather than intention to treat; the distribution of known confounders in the different treatment groups was not described; or the distribution of known confounders differed between the treatment groups but was not taken into account in the analyses. In nonrandomized studies, if the effect of the main confounders was not investigated or confounding was demonstrated but no adjustment was made in the final analyses, the question should be answered as “no”. | Yes = 1;<br>No = 0;<br>Unable to determine = 0 |
| **18. Did the authors present a power calculation including determining sample size necessary to detect relevant effect sizes?** | Yes = 1;<br>No = 0;<br>Unable to determine = 0 |

Adaptation of: Downs SH, Black N., The feasibility of creating a checklist for the assessment of the methodological quality both of randomised and non-randomised studies of health care interventions. J Epidemiol Community Health. 1998;52(6):377-84.

